# Exploring the impact of health expenditure and its allocation on neonatal and child mortality at national level across 188 countries from 2000 to 2019: insights from the Global Burden of Disease Study

**DOI:** 10.1101/2024.02.29.24303584

**Authors:** Ali Sheidaei, Negar Rezaei, Maryam Sharafkhah, Hossein Poustchi, Mohammadreza Mobinizadeh, Marita Mohammadshahi, Mohsen Naghavi, Alireza Olyaeemanesh, Reza Malekzadeh, Alireza Delavari, Sadaf G. Sepanlou

**Author notes:** **Corresponding author:** (SGS). These authors contributed equally to this work (Ali Sheidaei, Negar Rezaei).

## Abstract

**Background:** Exploring the impact of national health expenditure and its allocation on neonate and child mortality can help policy makers implement strategies aimed at achieving target 3.2 of Sustainable Development Goals (SDGs). The aim of the current study is to explore the impact of selected indicators of national health accounts on neonate and under-5 mortality across 188 countries from 2000 to 2019.

**Methods and findings:** This study has an ecological design. Data on health expenditure was obtained from the Global Health Expenditure Database (GHED) for 188 countries from 2000 to 2019. The Global Burden of Disease study (GBD) 2019 data on neonatal and under 5 mortality rates at national levels from 2000 to 2019 were obtained from the website of the Global Health Data Exchange (GHDx) supported by the Institute for Health Metrics and Evaluation. The income groups were stratified based on the World Bank classification. We employed a mixed-effects regression model to investigate the association of different health account indicators with changes in neonatal and under-5 mortality rates over time across countries. We used the Multiple Change Points model to determine the turning points in the association of health expenditure per capita with mortality across countries in 2019. And finally, we estimated the observed-to-expected ratio of mortality based on the segmented regression model for all 188 countries in 2019. Increase in the current health expenditure in International dollar Purchasing Power Parity (Int$ PPP) per capita was associated with lower mortality among both neonates and children in all strata of countries. Reductions were very minimal among high-income countries and were generally more prominent in low-income countries and decreased along with increase in income. Reductions were more noteworthy for under-5 mortality rates. The percentage of domestic general government health expenditure and the percentage of compulsory financing arrangements out of current health expenditure were inversely associated with mortality, while the association of percentage of domestic private health expenditure and out-of-pocket expenditure out of current health expenditure with mortality was positive. Results showed that the reduction in neonatal mortality associated with each ten-dollar increase in current health expenditure per capita is significantly more prominent for per capita expenditures less that the cut-point of 480 Int$ PPP per capita. The respective figure for under-5 mortality was 386 Int$ PPP per capita. Ultimately, a total of 110 countries had observed versus expected ratio less than one for neonatal mortality and 118 countries for child mortality.

**Conclusions:** Increase in health expenditure is significantly associated with decrease in neonate and under-5 mortality especially among low and low-middle income countries. However, the association fades among countries in which health expenditure per capita is higher than the threshold. In all countries, improvement in neonate and under-5 mortality requires modifying the health system infrastructure to move towards universal health coverage. However, the COVID-19 pandemic may have influenced the health spending at national levels.

## Introduction

Neonatal and child mortality have been considered as important indicators of countries’ overall health and development. The neonatal period is the most vulnerable time for a child’s survival. The Sustainable Development Goals (SDGs) aim to reduce neonatal mortality to at least as low as 12 deaths per 1,000 live births and under-5 mortality to at least as low as 25 deaths per 1,000 live births and to end preventable deaths in this age group by 2030 in all countries.[1] According to the SDGs Report 2022, crises have negatively affected the agenda for SDGs and threatened the human survival.[2] However, the world has made substantial progress in neonate and child survival since 1990, which has been least affected by the COVID-19 pandemic and the devastating conflicts[2] though there are studies showing that decline in healthcare utilization may reverse gains in reducing maternal and child mortality.[3] Globally, the number of neonatal deaths declined from 5 million in 1990 to 2.4 million in 2020.[4] In spite of this progress, further action is needed to meet the SDG targets and to ensure that every neonate and child has the chance to survive.

The association between health expenditure and neonate and child mortality is complex and varies across different regions and types of health expenditures.[5] Neonate and child mortality have several determinants such as poverty, education, urbanization, poor basic sanitation, and access to clean energy.[6–8] Access to health services for women and children, including prenatal care, skilled birth assistance, vaccination, and adequate nutrition are other determinants for neonate and child mortality.[9] Diseases such as HIV and malaria can negatively impact neonate and child survival.[10] Proper management of health care expenditure can positively impact this network of determinants. A more comprehensive overview of the impact of health expenditure and its allocation may help nations address child mortality.

Studies exploring the impact of health expenditure on neonate and child mortality date back to 1990s.[11] However, the impact is not straightforward, as it depends on how efficiently and equitably health resources are used, as well as on other determinants.[11] But thus far, the exact nature of the relationship, especially for public health expenditure, is not settled and results are controversial.[12–15] There is no consensus on the robust methods for exploring this association either. Some studies detected no association.[16, 17] However, other studies have found that the effect of health expenditure on infant mortality varies by income level of countries.[18, 19] For example, one study found that there is a clear positive and significant association between health expenditure and infant mortality only among high-income countries, while for low-income, lower-middle-income, and upper-middle-income countries, health spending does not have a significant association with infant health status.[20] The gap exists on effectiveness of health expenditure in itself in addition to types and sources of expenditure on mortality within and between countries among which, the health system infrastructure may vary across income levels.

There is additional controversy regarding the minimum required health expenditure to prevent neonatal and child mortality. There is no agreed-upon threshold for health expenditure per capita required to achieve the SDG targets for neonate and child mortality. Additionally, there is no metric to compare mortality between countries with comparable health expenditure per capita and to measure the exact performance of nations relative to the budget they spend on their health. Such information can have valuable policy implications and the potential to make those policies more effective and cost-effective at national level for all countries, globally.

Having said all challenges, the aim of this study is to examine the association between various health expenditure indicators and neonatal and child mortality across 188 countries from 2000 to 2019 and to develop a method for quantifying the performance of countries.

## Methods

### Study design and data sources

The current study has an ecological design. Data on health expenditure (National Health Accounts indicators) were obtained from the Global Health Expenditure Database (GHED) for 188 countries from 2000 to 2019.[21] The Global Burden of Disease study (GBD) 2019 data on neonatal and under 5 mortality rates at national levels from 2000 to 2019 were obtained from the website of the Global Health Data Exchange (GHDx) supported by the Institute for Health Metrics and Evaluation (IHME), which is an independent global health research center at the University of Washington. Data on income level of 188 countries were obtained from World Bank website. Data from the three sources were publicly available. We harmonized and merged the data from these sources.

### National Health Accounts (NHA) indicators

The main NHA indicators used in this study are defined as follows.

Current Health Expenditure (CHE) per capita is defined as the current expenditures on health per capita expressed in international dollars at purchasing power parity (PPP) to facilitate international comparisons. This indicator contributes to understanding the total expenditure on health relative to the beneficiary population. Domestic General Government Health Expenditure (GGHE-D) per capita is the part of CHE per capita provided by the domestic sources of the government. Domestic Private Health Expenditure (PVT-D) per capita is another indicator. This indicator calculates the average expenditures on health per capita funded by corporations and non-profit organizations. Out-of-Pocket Expenditure (OOPS) per capita are the costs of medical care that people pay for on their own and that are not covered or reimbursed by their insurance. They include deductibles, copays, coinsurance, and any services that are not covered by their health plan.

The rest of the indicators include the percentage of CHE that is allocated through the government (GGHE-D) or the private sector (PVT-D), or is directly paid by households (OOP). Government financing arrangements (GFA) are financing schemes, organized at a national level or for specific population groups that automatically entitle individuals to care based on residency, and form the principle mechanism by which health care expenses are covered in a number of countries. Finally, compulsory financing arrangements (CFA) are health insurance schemes (either through public or private entities) that are linked to the payment of social contributions or health insurance premiums and finance the bulk of health spending. And the last indicator is the proportion of gross domestic product (GDP) that is spent on health (CHE).

### Global Burden of Disease data

The detailed methodology for neonate and child mortality estimations in GBD have been previously published.[22] In short, for the estimation of neonate and child mortality in GBD, data were used from vital registration (VR) systems, sample registration systems (SRS), and disease surveillance point systems, household surveys (complete birth histories [CBH], summary birth histories [SBH]), censuses (SBHs, CBH in rare occasions), and Demographic Surveillance Sites (DSS). Livebirths and population in the under-5 age groups were produced as part of the GBD fertility and population estimation.

### World Bank Data

The World Bank classifies economies into four income groups: low income, lower-middle income, upper-middle income, and high income. The classification is based on gross national income (GNI) per capita data in U.S. dollars, converted from local currency using the World Bank Atlas method. The GNI per capita thresholds for 2019 are: low income: less than $996, lower-middle income: between $996 and $3,895, upper-middle income: between $3,896 and $12,055, high income: greater than $12,055. The classification is updated annually based on the estimate of GNI per capita for the previous calendar year. The estimates of GNI are obtained from economists in World Bank country units who rely primarily on official data published by the countries.

### Statistical methods

In order to investigate the association of different health account indicators with changes in neonatal and under-5 mortality rates over time across countries with various income, we employed a mixed-effects regression model. Our initial analyses on countries stratified into four income levels demonstrated substantial heterogeneity in mortality rate within strata, which led to unstable estimations. Thus, we further divided countries into low mortality and high mortality. We considered specific thresholds of 12 deaths per 1,000 live births for the neonatal mortality rate and 25 deaths per 1,000 live births for the under-5 mortality rate, in line with SDG Target 3.2. Thus, the model was stratified based on four income levels and two levels of mortality rates, ending in eight sub-groups. However, instead of eight strata, this categorization resulted in a total of six strata as there was no low income country with low mortality and no high income country with high mortality. Stratification significantly reduced the variance of estimates. Within each stratum, the model included the fixed effect of the health account indicator and a random effect for each country as predictors:

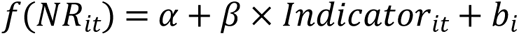

Here, *NR*_*it*_ represents the neonatal or under-5 mortality rate for country *i* in year *t*, *f*(*x*) denotes the link function (both identity and logarithm were considered), and *b*_*i*_ is the random effect for country i, following a 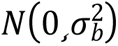 distribution.

Subsequently, we focused on year 2019, where we examined variations across countries. To accomplish this, we employed a Bayesian inference approach known as Multiple Change Points (MCP) between linear segments. To determine the optimal number of linear segmentations, we fitted a linear model with a single segmentation and compared it with models containing 2, 3, and 4 segmentations. The Akaike Information Criterion (AIC) and Bayesian Information Criterion (BIC) were utilized to compare the models and identify the most suitable number of thresholds.

To ensure the robustness of our findings, we conducted a sensitivity analysis. Firstly, we repeated the MCP approach for all countries, regardless of income level, and estimated the observed-to-expected ratio of neonatal and under-5 mortalities. Additionally, we performed the analysis by excluding high-income countries and again estimated the observed-to-expected ratio of mortalities. Analyses were performed in Stata software V.14 (StataCorp, College Station, TX, USA) and R-4.2.2.

## Results

### Mixed-effects linear models

Six strata were defined based on income level and mortality rate. Figure 1 shows the classification of countries into the strata. Table 1 demonstrates the coefficients from the mixed effects model. The results are consistent for both neonatal mortality and under-5 mortality. Indicators representing per capita expenditure, including current health expenditure per capita, domestic general government health expenditure per capita, domestic private health expenditure per capita, and out-of-pocket expenditure per capita were inversely associated with neonatal and under-5 mortality per 1 000 live births (Table 1). According to this assessment, each ten international dollar increase in current health expenditure per capita was associated with a reduction of −0.19 (95% CI: −0.21, −0.18) neonatal mortality per 1,000 live births in low-middle income countries with low mortality, and a reduction of −0.05 (−0.06, −0.05) in up-middle income countries with low mortality. The ten international dollar increase in current health expenditure per capita was also associated with a reduction of −0.65 (−0.73, −0.57) neonatal mortality rate per 1,000 live births in low-income countries with high mortality. Respective estimates for low-middle income and up-middle income countries with high mortality were −0.35 (−0.39, −0.31) and −0.11 (−0.13, −0.1). Reductions were very minimal among high-income countries. Generally, reductions were more prominent in low-income countries and decreased along with increase in income.

**Fig 1.**
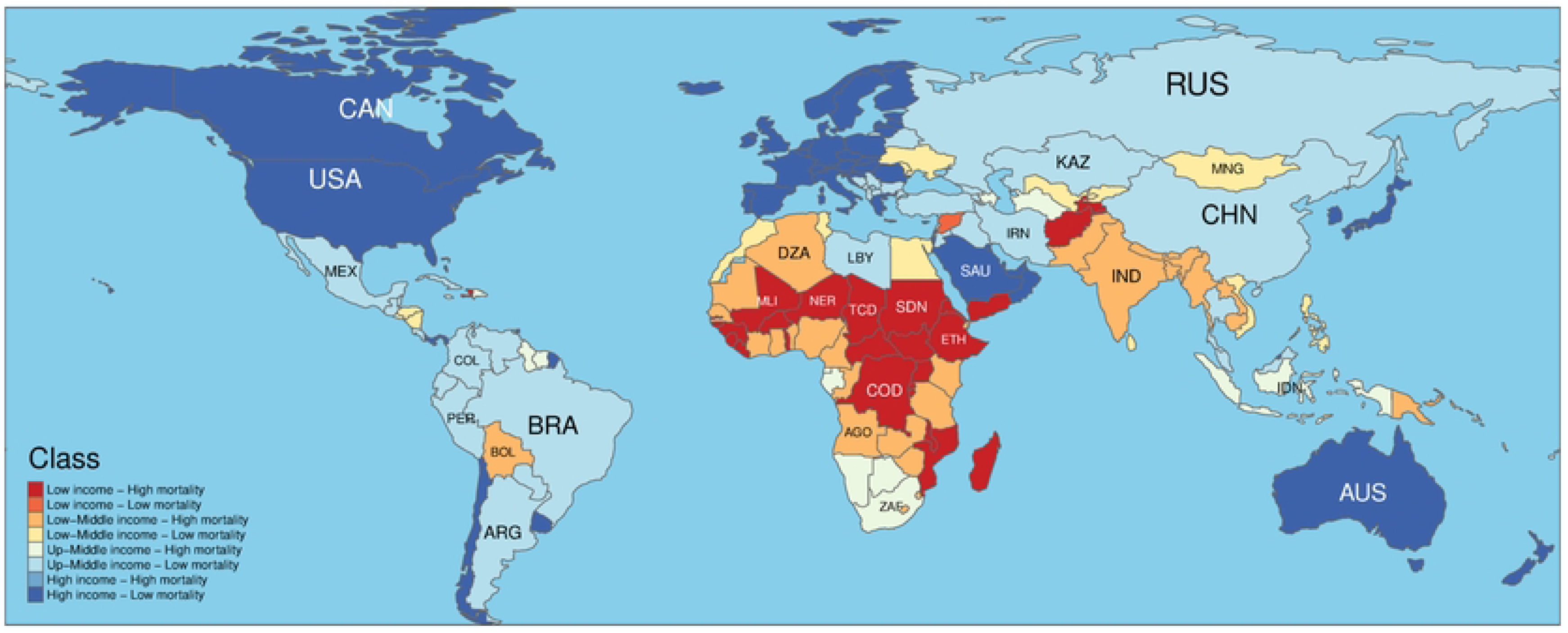
Classification of countries into six strata.

**Table 1.** Coefficients of the random effects model on association of each of indicators presenting per capita expenditure with neonatal and child mortality.

| Neonatal Mortality |  |  |  |  |  |
| --- | --- | --- | --- | --- | --- |
| Indicator* | Mortality level | Income level |  |  |  |
|  |  | Low | Low-Middle | Up-Middle | High |
| CHE per capita | Low | - | -0.19 (-0.21,-0.18) | -0.05 (-0.06,-0.05) | -0.01 (-0.01,0) |
| GGHED per capita | Low | - | -0.34 (-0.38,-0.3) | -0.06 (-0.07,-0.06) | -0.01 (-0.01,-0.01) |
| PVTD per capita | Low | - | -0.37 (-0.41,-0.33) | -0.12 (-0.13,-0.11) | -0.01 (-0.02,-0.01) |
| OOP per capita | Low | - | -0.38 (-0.43,-0.34) | -0.12 (-0.13,-0.1) | -0.03 (-0.03,-0.02) |
| CHE per capita | High | -0.65 (-0.73,-0.57) | -0.35 (-0.39,-0.31) | -0.11 (-0.13,-0.1) | - |
| GGHED per capita | High | -1.26 (-1.64,-0.88) | -0.44 (-0.51,-0.37) | -0.14 (-0.17,-0.12) | - |
| PVTD per capita | High | -1.02 (-1.16,-0.87) | -0.92 (-1.03,-0.81) | -0.18 (-0.21,-0.16) | - |
| OOP per capita | High | -0.99 (-1.14,-0.83) | -0.9 (-1.02,-0.77) | -0.18 (-0.22,-0.15) | - |
| Child Mortality |  |  |  |  |  |
| Indicator* | Mortality level | Income level |  |  |  |
|  |  | Low | Low-Middle | Up-Middle | High |
| CHE per capita | Low | - | -0.41 (-0.45,-0.37) | -0.11 (-0.12,-0.1) | -0.01 (-0.01,-0.01) |
| GGHED per capita | Low | - | -0.69 (-0.77,-0.61) | -0.13 (-0.15,-0.12) | -0.02 (-0.02,-0.01) |
| PVTD per capita | Low | - | -0.8 (-0.89,-0.71) | -0.25 (-0.27,-0.22) | -0.03 (-0.03,-0.02) |
| OOP per capita | Low | - | -0.84 (-0.94,-0.74) | -0.24 (-0.27,-0.21) | -0.06 (-0.06,-0.05) |
| CHE per capita | High | -3.7 (-4.17,-3.22) | -1.76 (-1.98,-1.55) | -0.4 (-0.47,-0.34) | - |
| GGHED per capita | High | -7.94 (-10.1,-5.8) | -2.19 (-2.58,-1.8) | -0.55 (-0.66,-0.44) | - |
| PVTD per capita | High | -5.16 (-6.01,-4.3) | -3.69 (-4.3,-3.06) | -0.62 (-0.75,-0.49) | - |
| OOP per capita | High | -4.88 (-5.79,-3.96) | -3.48 (-4.15,-2.79) | -0.55 (-0.71,-0.39) | - |

Per capita expenditures are reported in purchasing power parity of international dollar
\*Indicators: CHE= Current Health Expenditure. GGHED= Domestic General Government Health Expenditure. PVTD=Domestic Private Health Expenditure. OOP= Out-of-Pocket Expenditure.

Reductions were more noteworthy for under-5 mortality rates (Table 1). Each ten international dollar increase in current health expenditure per capita was associated with a reduction of −0.41 (−0.45, −0.37) in child mortality per 1,000 live births in low-middle income countries with low mortality, and a reduction of −0.11 (−0.12, −0.1) in up-middle income countries with low mortality. The ten international dollar increase in current health expenditure per capita was also associated with a reduction of −3.7 (−4.17, −3.22) in child mortality rate per 1,000 live births in low-income countries with high mortality. Respective estimates for low-middle income and up-middle income countries with high mortality were −1.76 (−1.98, −1.55) and −0.4 (−0.47, −0.34). Again, reductions were very minimal among high-income countries and reductions were more prominent in low-income countries and decreased along with increase in income. Reductions were also more prominent in countries with high mortality compared with their counterparts with low mortality. Other indicators per capita showed quite similar patterns, even for PVTD per capita and OOP per capita (Table 1).

The association between the percentage of domestic general government health expenditure out of current health expenditure with mortality was inverse in all strata except for the low-income countries with high mortality (Table 2). However, one percent increase in the percentage of domestic private health expenditure and out-of-pocket expenditure out of current health expenditure was associated with higher mortality rates for both neonates and children under 5 years in all categories (Table 2). The association of percentage of government financing arrangements out of current health expenditure was not consistent across six categories of countries. However, the percentage of compulsory financing arrangements out of current health expenditure was consistently associated with lower mortality in all six categories and for both neonates and under-5 children. The association of percentage of current health expenditure out of gross domestic product with mortality was also inverse in all categories except for low-middle income countries with high mortality [20.27 (4.43, 35.65)].

**Table 2.**
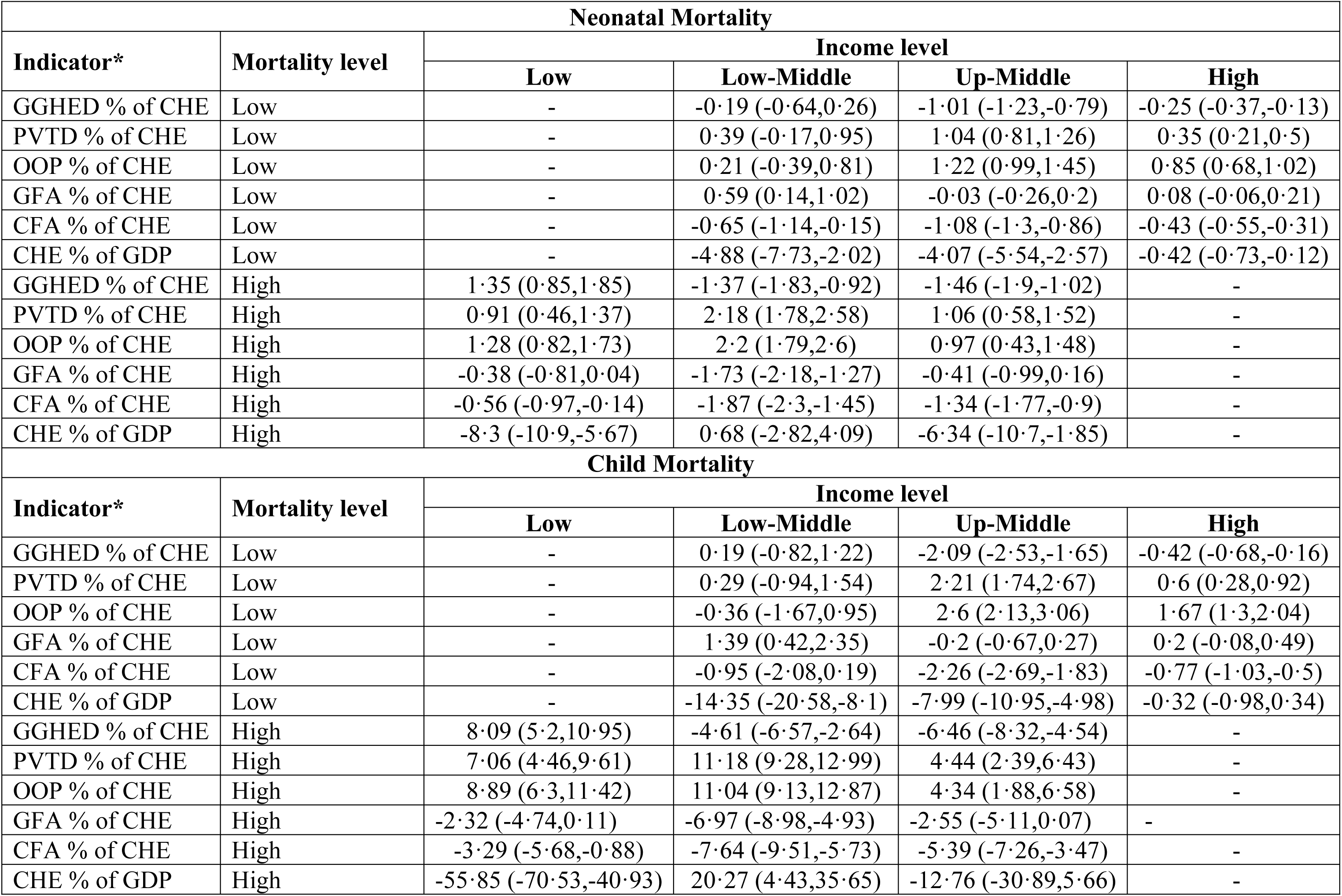
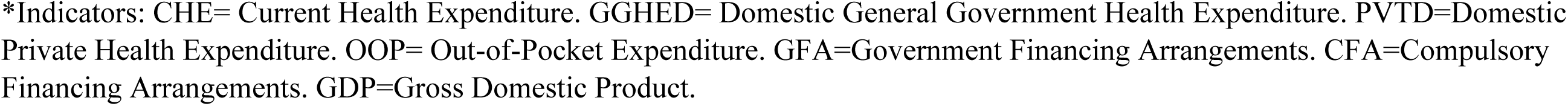
Coefficients of the random effects model on association of percentage indicators with neonatal and under-5 mortality.

| Neonatal Mortality |  |  |  |  |  |
| --- | --- | --- | --- | --- | --- |
| Indicator* | Mortality level | Income level |  |  |  |
|  |  | Low | Low-Middle | Up-Middle | High |
| GGHED % of CHE | Low | - | -0.19 (-0.64,0.26) | -1.01 (-1.23,-0.79) | -0.25 (-0.37,-0.13) |
| PVTD % of CHE | Low | - | 0.39 (-0.17,0.95) | 1.04 (0.81,1.26) | 0.35 (0.21,0.5) |
| OOP % of CHE | Low | - | 0.21 (-0.39,0.81) | 1.22 (0.99,1.45) | 0.85 (0.68,1.02) |
| GFA % of CHE | Low | - | 0.59 (0.14,1.02) | -0.03 (-0.26,0.2) | 0.08 (-0.06,0.21) |
| CFA % of CHE | Low | - | -0.65 (-1.14,-0.15) | -1.08 (-1.3,-0.86) | -0.43 (-0.55,-0.31) |
| CHE % of GDP | Low | - | -4.88 (-7.73,-2.02) | -4.07 (-5.54,-2.57) | -0.42 (-0.73,-0.12) |
| GGHED % of CHE | High | 1.35 (0.85,1.85) | -1.37 (-1.83,-0.92) | -1.46 (-1.9,-1.02) | - |
| PVTD % of CHE | High | 0.91 (0.46,1.37) | 2.18 (1.78,2.58) | 1.06 (0.58,1.52) | - |
| OOP % of CHE | High | 1.28 (0.82,1.73) | 2.2 (1.79,2.6) | 0.97 (0.43,1.48) | - |
| GFA % of CHE | High | -0.38 (-0.81,0.04) | -1.73 (-2.18,-1.27) | -0.41 (-0.99,0.16) | - |
| CFA % of CHE | High | -0.56 (-0.97,-0.14) | -1.87 (-2.3,-1.45) | -1.34 (-1.77,-0.9) | - |
| CHE % of GDP | High | -8.3 (-10.9,-5.67) | 0.68 (-2.82,4.09) | -6.34 (-10.7,-1.85) | - |
| Child Mortality |  |  |  |  |  |
| Indicator* | Mortality level | Income level |  |  |  |
|  |  | Low | Low-Middle | Up-Middle | High |
| GGHED % of CHE | Low | - | 0.19 (-0.82,1.22) | -2.09 (-2.53,-1.65) | -0.42 (-0.68,-0.16) |
| PVTD % of CHE | Low | - | 0.29 (-0.94,1.54) | 2.21 (1.74,2.67) | 0.6 (0.28,0.92) |
| OOP % of CHE | Low | - | -0.36 (-1.67,0.95) | 2.6 (2.13,3.06) | 1.67 (1.3,2.04) |
| GFA % of CHE | Low | - | 1.39 (0.42,2.35) | -0.2 (-0.67,0.27) | 0.2 (-0.08,0.49) |
| CFA % of CHE | Low | - | -0.95 (-2.08,0.19) | -2.26 (-2.69,-1.83) | -0.77 (-1.03,-0.5) |
| CHE % of GDP | Low | - | -14.35 (-20.58,-8.1) | -7.99 (-10.95,-4.98) | -0.32 (-0.98,0.34) |
| GGHED % of CHE | High | 8.09 (5.2,10.95) | -4.61 (-6.57,-2.64) | -6.46 (-8.32,-4.54) | - |
| PVTD % of CHE | High | 7.06 (4.46,9.61) | 11.18 (9.28,12.99) | 4.44 (2.39,6.43) | - |
| OOP % of CHE | High | 8.89 (6.3,11.42) | 11.04 (9.13,12.87) | 4.34 (1.88,6.58) | - |
| GFA % of CHE | High | -2.32 (-4.74,0.11) | -6.97 (-8.98,-4.93) | -2.55 (-5.11,0.07) | - |
| CFA % of CHE | High | -3.29 (-5.68,-0.88) | -7.64 (-9.51,-5.73) | -5.39 (-7.26,-3.47) | - |
| CHE % of GDP | High | -55.85 (-70.53,-40.93) | 20.27 (4.43,35.65) | -12.76 (-30.89,5.66) | - |

\*Indicators: CHE= Current Health Expenditure. GGHE= Domestic General Government Health Expenditure. PVT=Domestic Private Health Expenditure. OOP= Out-of-Pocket Expenditure. GFA=Government Financing Arrangements. CFA=Compulsory Financing Arrangements. GDP=Gross Domestic Product.

### Multiple change points models

The output of multiple change points is presented in Table 3 and Figure 2. Estimates showed that the slope of the inverse association between per capita expenditure and mortality is significantly different before and after the change point. Results showed that the reduction in neonatal mortality associated with each ten-dollar increase in current health expenditure per capita is significantly more prominent for per capita expenditures less that the cut-point of 480 International dollars. The decrease in neonatal mortality with each ten-dollar increase is much less when the current health expenditure per capita is more than 480 International dollars. The same pattern is observed for under 5 mortality where the cut-point for current health expenditure per capita is 386 International dollars. The same pattern is also observed for health expenditure in other sectors of the health system: the general government health expenditure, the domestic private health expenditure, and the out-of-pocket health expenditure (supplement). In the sensitivity analyses we excluded high-income countries from the model and again estimated the cut-points. Slopes before and after the change point were similar. Yet, the cut-offs were relatively lower (Table 3).

**Fig 2.**
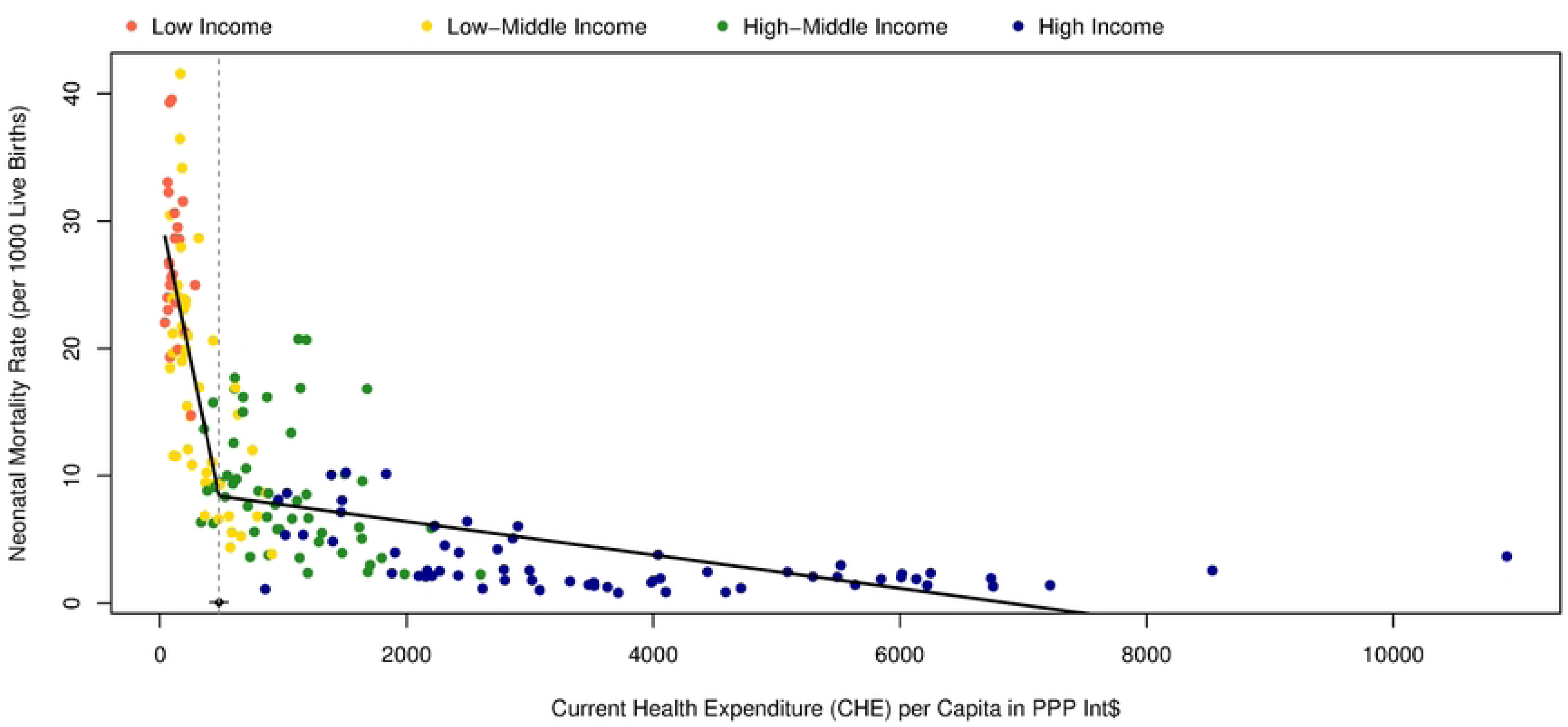
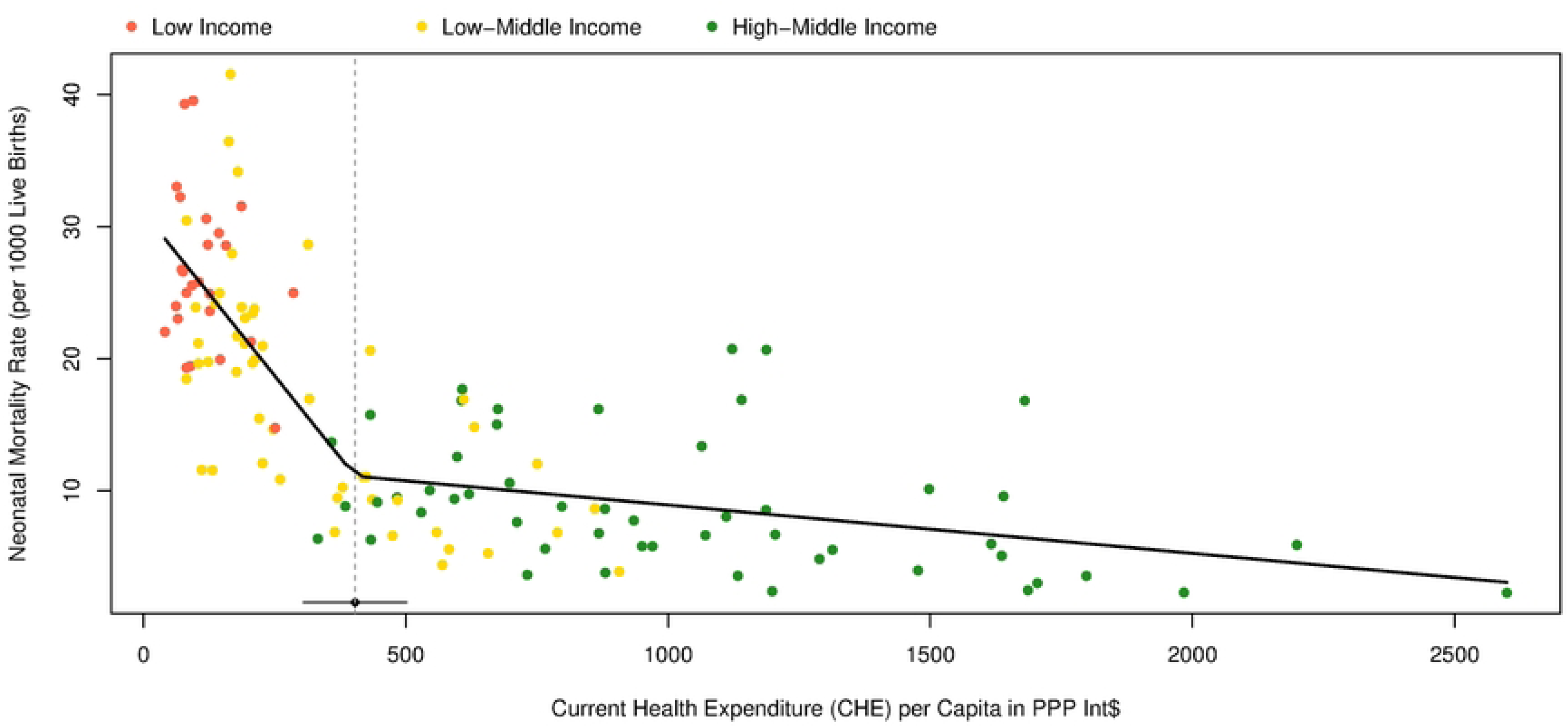
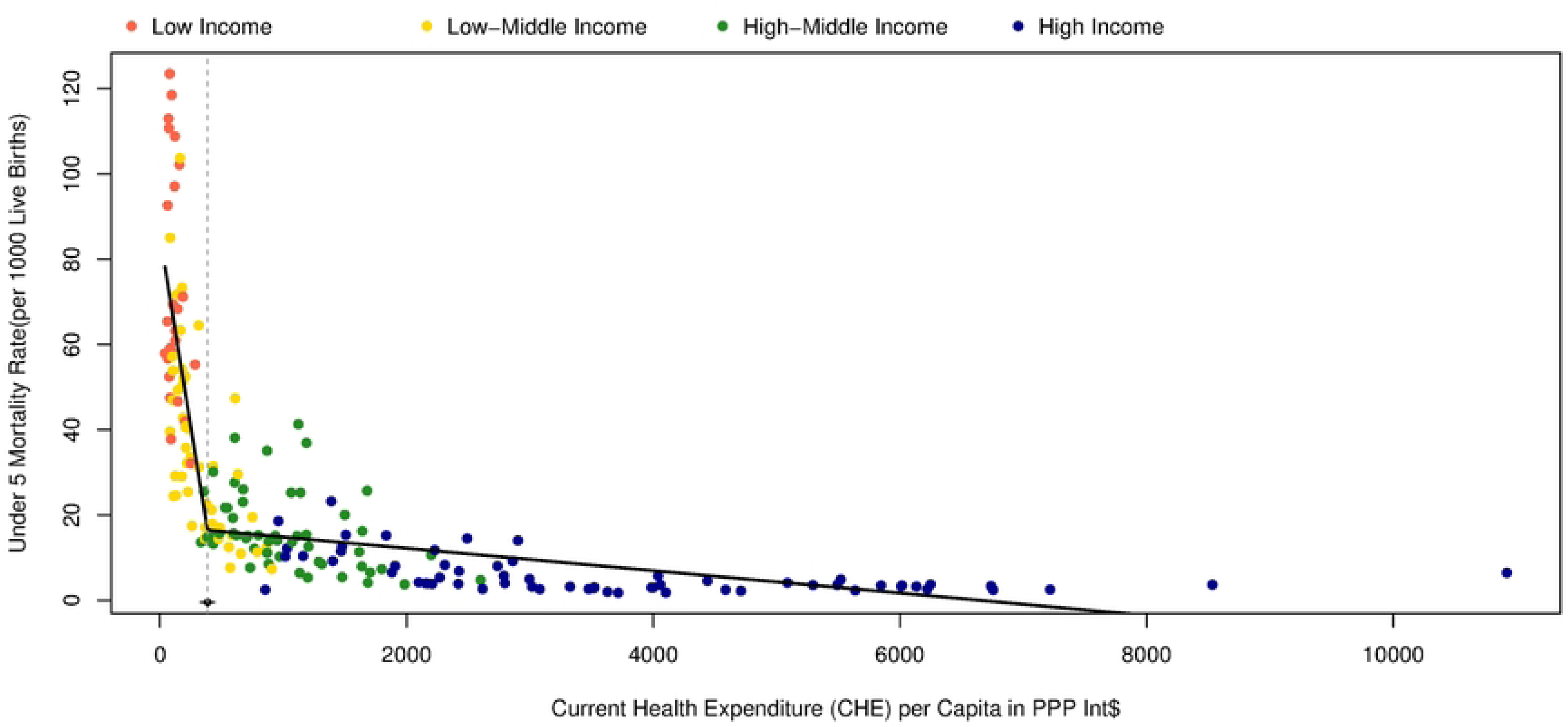
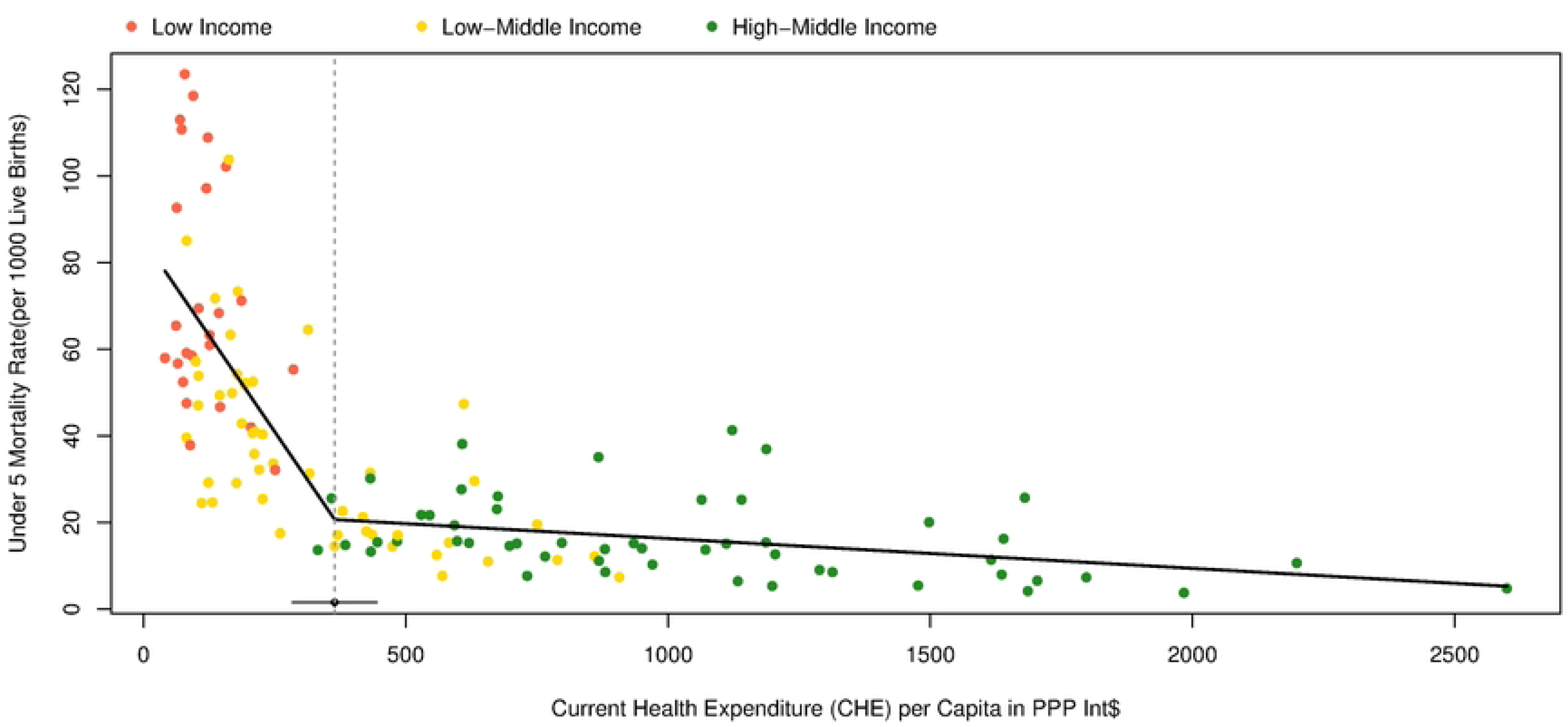
The results of the multiple change points models for the association of current per capita expenditure with mortality in 2019. A) Neonatal mortality in all countries; B) Neonatal mortality excluding high-income countries; C) Under-5 mortality in all countries; D) Under-5 mortality excluding high-income countries.

**Table 3.**
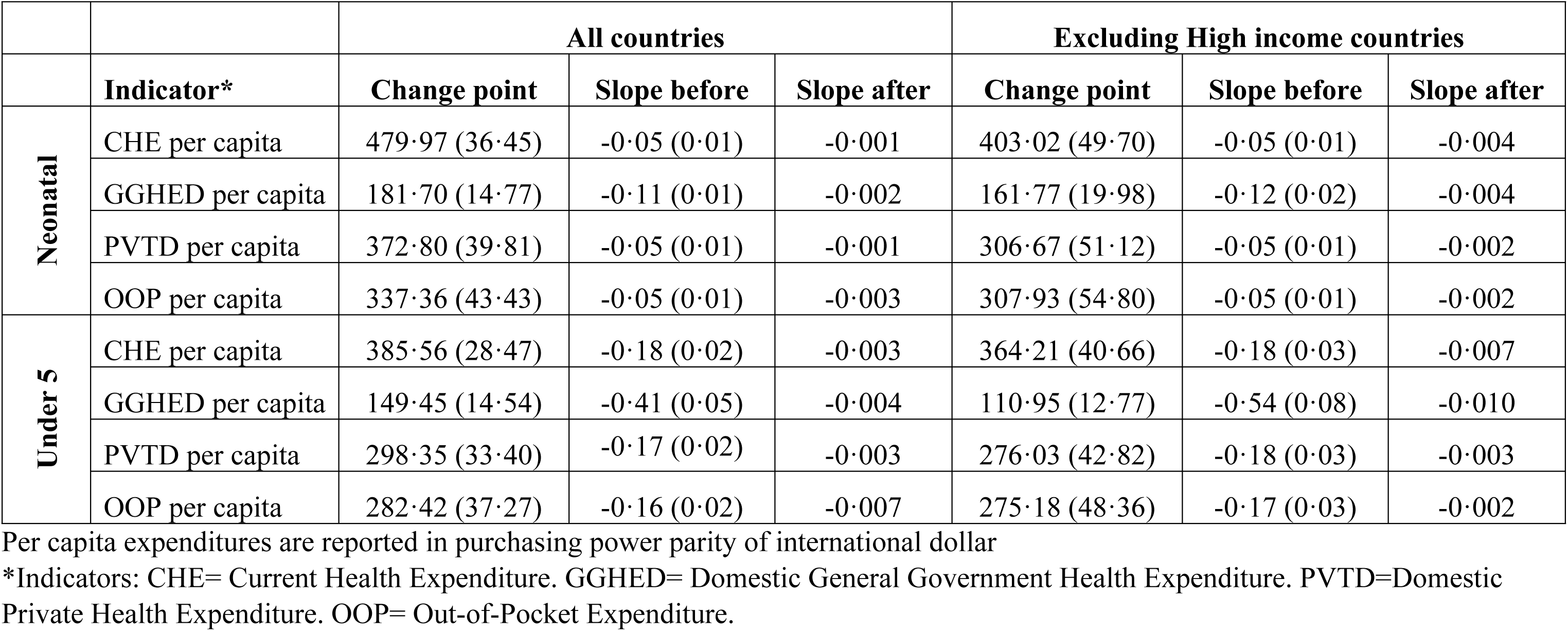
Output of the multiple change points for per capita indicators. Results are presented for the main analysis including all countries and the sensitivity analysis excluding high-income countries.

Supplementary tables (Stables 1 to 8) and Figure 3 show the expected neonate and under 5 mortality rates based on per capita health expenditures and the observed versus expected mortality for 188 countries in 2019. A total of 110 countries had observed versus expected ratio less than one for neonatal mortality and 118 countries for child mortality. The United States has the largest current health expenditure per capita but its observed neonatal mortality and the under 5 mortality are much higher than the expected based on current health expenditure per capita. The demographic republic of Congo has the lowest current health expenditure per capita but the observed versus expected neonatal and under 5 mortality in this country are 0.77 and 0.74 respectively. Germany, the Netherlands, Luxemburg, Norway, Switzerland, and the United States are high-income countries which have a higher observed versus expected neonatal and under-5 mortality based on per capita current health expenditure. Cook Islands and Andorra are high-income countries that have the lowest observed versus expected neonatal and under-5 mortality rates (Figure 3). Among low-income countries, Eritrea (0.72) and Gambia (0.73) had the lowest observed versus expected ratio for neonatal mortality and Mali (1.51) and Central African Republic (1.46) had highest observed versus expected ratio. Respective figures for under-5 mortality were Gambia (0.54) and Eritrea (0.67) with lowest ratio and Sierra Leone (1.78), Mali (1.73), and Central African Republic (1.73) with highest ratio. Among low-middle income countries, Vanuatu had the lowest observed versus expected ratio for both neonatal (0.45) and child (0.37) mortality. The highest observed to expected ratio among low-middle income countries belonged to Eswatini, with the ratio of 2.06 for neonatal mortality and 2.99 for child mortality. Finally, among upper-middle income group, Belarus, Montenegro, and Serbia had the lowest observed versus expected ratio and South Africa and Botswana had the highest ratio for both neonatal and under-5 mortality.

**Fig 3.**
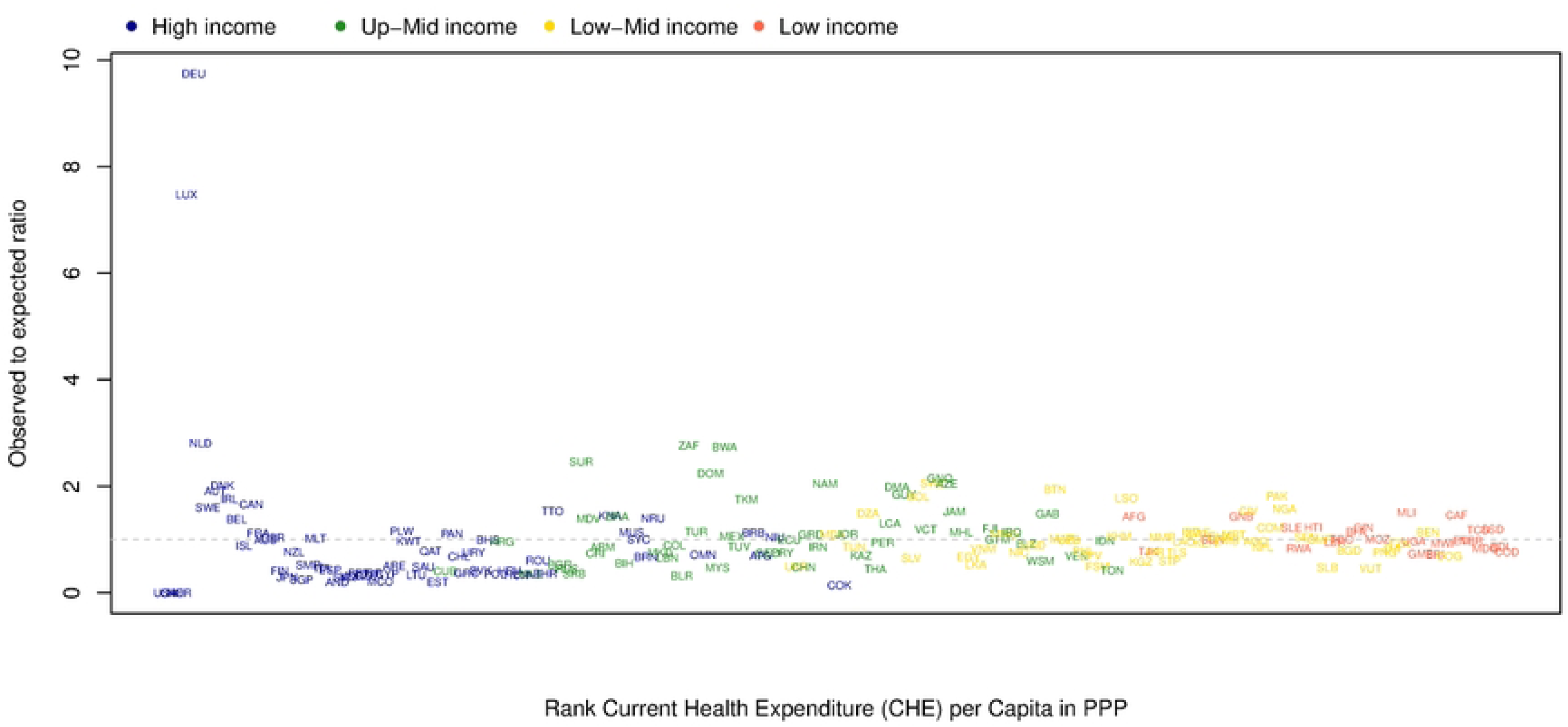
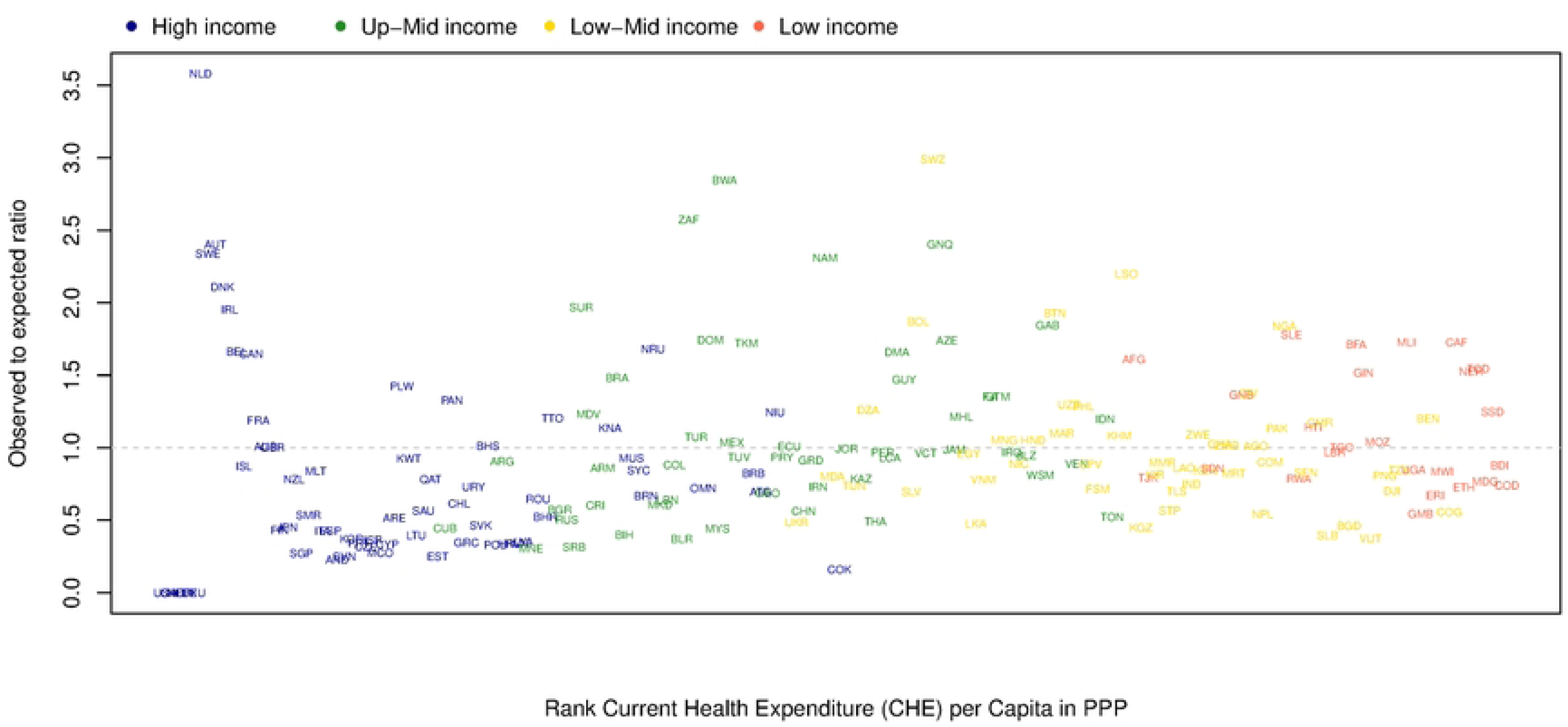
Output of the multiple change points for the observed versus expected ratio of mortality based on current health expenditure per capita (Int$) among 188 countries in 2019. A) Neonatal mortality; B) Under-5 mortality.

## Discussion

To put it in a nutshell, the results of our study showed that higher health expenditure per capita is associated with a lower mortality among neonates and children. Additionally, along with increase in percentage of government health expenditure and compulsory financial arrangements out of the total current health expenditure, mortality decreases. However, any increase in percentage of private health expenditure and out-of-pocket health expenditure out of the total current health expenditure are associated with higher mortality among both neonates and children.

Our results demonstrated that increase in per capita expenditure is more effective in decreasing mortality among low-income countries. Additionally, among countries with low-middle or upper-middle income, increase in expenditure per capita is more prominent in high mortality countries compared to their low mortality counterparts. Our results also demonstrated that there is a cut-off in per capita expenditure below which increase in per capita expenditure significantly decreases the neonate and child mortality and above which, the decrease in mortality becomes non-significant. This observation shows that above the cut-off, increase in expenditure has no added value if the infrastructure of the system is not optimized. It shows that among all countries, regardless of their income level, increase in the share of domestic general government expenditure out of current health expenditure is associated with decrease in mortality. However, the increase in share of domestic private health expenditure and out-of-pocket expenditure increases mortality. Another finding was that the share of government financing arrangements out of current health expenditure had no consistent association with mortality across countries. However, the association for compulsory financing arrangements was inverse across all categories of countries.

As mentioned before, previous studies present contradictory results on association of various health expenditure indicators with neonate and child mortality. The empirical results of a study in India showed that publicly financed health expenditure reduces infant mortality, child mortality.[23] Another study utilizing panel data from 2000 to 2015 across 46 countries in Sub-Saharan Africa found that total health expenditure, which includes public, private, and external expenditures, significantly impacts infant and neonatal mortality rates. This study indicates that higher health expenditures are generally associated with lower mortality rates, but the impact varies when considering public, private, and external sources of health funding separately.[24] Both public and external sources of health expenditure had a significant negative association with neonate and infant mortality but private association had no significant association.[24] This is in contrast with our study results which show health expenditure per capita regardless of the source is associated with lower mortality specially in low and low-middle income countries. Consistent with our results, other studies showed that reduced user charges were associated with improved health outcomes, particularly for lower-income groups and children in low and middle income countries.[25]

We found more prominent associations between health expenditure and under 5 mortality, which is consistent with a number of previous reports.[26] However, There are other studies that show differential impact of health expenditure on children at different age groups.[16] A recent study by Garcia et al reported a negative association between total health expenditure and specifically public expenditure with neonate mortality[27] which aligns with previous studies.[5, 16, 18, 19, 28, 29] However, the association appeared to be non-significant for under 5 mortality[27], which is inconsistent with our results. We couldn’t find any justification for the inconsistency.

These results, collectively imply that enhancing universal health coverage using any strategy, no matter through public or private venues, can effectively decrease neonate and under-5 mortality. Any progress towards universal health coverage through prepayment mechanisms such as insurance and taxation can improve health outcomes and reduce health inequalities in all countries.[25]

The main limitation of our study is its ecological design. Every country has its own health system infrastructure, with the interplay between sources of funds through the government, private sector, donors, and households, which is affected by the overall level of health expenditure at national levels, the composition of the population (age structure, urbanization, education, sanitation, immunization), and the extent and share of primary health care in the overall health system of the country. Unfortunately, data on the overall expenditure on primary health care was not available for most of the countries, which made it impossible to examine its impact on mortality. Despite all of these limitations, we found significant results on the role of various sources of funds at national level on neonate and child mortality with important policy implications for policy makers at national and even provincial level for 188 countries and territories globally.

## Conclusions

Our results highlight the necessity of achieving universal health coverage including financial risk protection, as one of the main SDG targets by 2030. Increase in per capita is effective in countries with low expenditure on health. However, above the threshold, it is recommended that the infrastructure of the system be optimized to achieve further progress in lowering neonatal and under-5 mortality. Policy makers in high mortality nations can take advantage of their low mortality counterparts to improve the effectiveness and cost-effectiveness of their health system.

## Data Availability

All of the data are publicly available in the site of the World Health Organization, the World Bank, and the Institute for Health Metrics and Evaluation.

https://data.worldbank.org/country

https://vizhub.healthdata.org/gbd-results/

https://apps.who.int/nha/database/Select/Indicators/en

## Acknowledgements

None.

## Supporting information

**S1 Fig. The results of the multiple change points models for the association of Domestic General Government Health Expenditure per capita expenditure with neonatal mortality in 2019**

**S2 Fig. The results of the multiple change points models for the association of Domestic General Government Health Expenditure per capita expenditure with neonatal mortality in 2019 excluding high-income countries**

**S3 Fig. The results of the multiple change points models for the association of Domestic General Government Health Expenditure per capita expenditure with under-5 mortality in 2019**

**S4 Fig. The results of the multiple change points models for the association of Domestic General Government Health Expenditure per capita expenditure with under-5 mortality in 2019 excluding high-income countries**

**S5 Fig. The results of the multiple change points models for the association of Domestic Private Health Expenditure per capita expenditure with neonatal mortality in 2019**

**S6 Fig. The results of the multiple change points models for the association of Domestic Private Health Expenditure per capita expenditure with neonatal mortality in 2019 excluding high-income countries.**

**S7 Fig. The results of the multiple change points models for the association of Domestic Private Health Expenditure per capita expenditure with under-5 mortality in 2019**

**S8 Fig. The results of the multiple change points models for the association of Domestic Private Health Expenditure per capita expenditure with under-5 mortality in 2019 excluding high-income countries**

**S9 Fig. The results of the multiple change points models for the association of Out−of−Pocket Expenditure per capita expenditure with neonatal mortality in 2019**

**S10 Fig. The results of the multiple change points models for the association of Out−of−Pocket Expenditure per capita expenditure with neonatal mortality in 2019 excluding high-income countries.**

**S11 Fig. The results of the multiple change points models for the association of Out−of−Pocket Expenditure per capita expenditure with under-5 mortality in 2019**

**S12 Fig. The results of the multiple change points models for the association of Out−of−Pocket Expenditure per capita expenditure with under-5 mortality in 2019 excluding high-income countries**

**S13 Fig. Output of the multiple change points for the observed versus expected ratio of neonatal mortality among 188 countries based on Domestic General Government Health Expenditure per capita in 2019**

**S14 Fig. Output of the multiple change points for the observed versus expected ratio of under-5 mortality among 188 countries based on Domestic General Government Health Expenditure per capita in 2019**

**S15 Fig. Output of the multiple change points for the observed versus expected ratio of neonatal mortality among 188 countries based on Domestic Private Health Expenditure per capita in 2019**

**S16 Fig. Output of the multiple change points for the observed versus expected ratio of under-5 mortality among 188 countries based on Domestic Private Health Expenditure per capita in 2019**

**S17 Fig. Output of the multiple change points for the observed versus expected ratio of neonatal mortality among 188 countries based on Out-of-Pocket Expenditure per capita in 2019**

**S18 Fig. Output of the multiple change points for the observed versus expected ratio of under-5 mortality among 188 countries based on Out-of-Pocket Expenditure per capita in 2019**

**S1 Table. Output of the multiple change points for observed versus expected neonatal mortality rate based on current health expenditure per capita (Int$) across 188 countries in 2019. For the observed versus expected in selected countries, high-income countries have been excluded from the model.**

**S2 Table. Output of the multiple change points for observed versus expected under 5 mortality rate based on current health expenditure per capita (Int$) across 188 countries in 2019. For the observed versus expected in selected countries, high-income countries have been excluded from the model.**

**S3 Table. Output of the multiple change points for observed versus expected neonatal mortality rate based on general government health expenditure per capita (Int$) across 188 countries in 2019. For the observed versus expected in selected countries, high-income countries have been excluded from the model.**

**S4 Table. Output of the multiple change points for observed versus expected under 5 mortality rate based on general government health expenditure per capita (Int$) across 188 countries in 2019. For the observed versus expected in selected countries, high-income countries have been excluded from the model.**

**S5 Table. Output of the multiple change points for observed versus expected neonatal mortality rate based on domestic private health expenditure per capita (Int$) across 188 countries in 2019. For the observed versus expected in selected countries, high-income countries have been excluded from the model.**

**S6 Table. Output of the multiple change points for observed versus expected under 5 mortality rate based on domestic private health expenditure per capita (Int$) across 188 countries in 2019. For the observed versus expected in selected countries, high-income countries have been excluded from the model.**

**S7 Table. Output of the multiple change points for observed versus expected neonatal mortality rate based on out-of-pocket expenditure per capita (Int$) across 188 countries in 2019. For the observed versus expected in selected countries, high-income countries have been excluded from the model.**

**S8 Table. Output of the multiple change points for observed versus expected under 5 mortality rate based on out-of-pocket expenditure per capita (Int$) across 188 countries in 2019. For the observed versus expected in selected countries, high-income countries have been excluded from the model.**

## References

1. Sustainable Development Goals. https://www.un.org/sustainabledevelopment (last accessed December 30, 2023).

2. United Nations. Sustainable Development Goals Report 2022. https://unstats.un.org/sdgs/report/2022/ (last accessed December 30th, 2023).

3. Ahmed T, Roberton T, Vergeer P, Hansen PM, Peters MA, Ofosu AA, et al. Healthcare utilization and maternal and child mortality during the COVID-19 pandemic in 18 low- and middle-income countries: An interrupted time-series analysis with mathematical modeling of administrative data. PLoS medicine. 2022;19(8):e1004070. Epub 2022/08/31. doi: 10.1371/journal.pmed.1004070. PubMed PMID: 36040910; PubMed Central PMCID: PMCPMC9426906.

4. Newborn Mortality, World Health Organization. https://www.who.int/news-room/fact-sheets/detail/levels-and-trends-in-child-mortality-report-2021 (last accessed July 24th, 2023).

5. Kipp AM, Blevins M, Haley CA, Mwinga K, Habimana P, Shepherd BE, et al. Factors associated with declining under-five mortality rates from 2000 to 2013: an ecological analysis of 46 African countries. BMJ open. 2016;6(1):e007675. Epub 2016/01/10. doi: 10.1136/bmjopen-2015-007675. PubMed PMID: 26747029; PubMed Central PMCID: PMCPMC4716228.

6. Cohen RL, Murray J, Jack S, Arscott-Mills S, Verardi V. Impact of multisectoral health determinants on child mortality 1980-2010: An analysis by country baseline mortality. PloS one. 2017;12(12):e0188762. Epub 2017/12/07. doi: 10.1371/journal.pone.0188762. PubMed PMID:29211765; PubMed Central PMCID: PMCPMC5718556.

7. Mohamoud YA, Kirby RS, Ehrenthal DB. Poverty, urban-rural classification and term infant mortality: a population-based multilevel analysis. BMC pregnancy and childbirth. 2019;19(1):40. Epub 2019/01/24. doi: 10.1186/s12884-019-2190-1. PubMed PMID: 30669972; PubMed Central PMCID: PMCPMC6343321.

8. Victora C, Boerma T, Requejo J, Mesenburg MA, Joseph G, Costa JC, et al. Analyses of inequalities in RMNCH: rising to the challenge of the SDGs. BMJ global health. 2019;4(Suppl 4):e001295. Epub 2019/07/13. doi: 10.1136/bmjgh-2018-001295. PubMed PMID: 31297251; PubMed Central PMCID: PMCPMC6590961.

9. Victora CG, Requejo JH, Barros AJ, Berman P, Bhutta Z, Boerma T, et al. Countdown to 2015: a decade of tracking progress for maternal, newborn, and child survival. Lancet (London, England). 2016;387(10032):2049–59. Epub 2015/10/20. doi: 10.1016/s0140-6736(15)00519-x. PubMed PMID: 26477328; PubMed Central PMCID: PMCPMC7613171.

10. Goga A, Singh Y, Jackson D, Pillay Y, Bhardwaj S, Chirinda W, et al. Is elimination of vertical transmission of HIV in high prevalence settings achievable? BMJ (Clinical research ed). 2019;364:l687. Epub 2019/04/09. doi: 10.1136/bmj.l687. PubMed PMID: 30957782; PubMed Central PMCID: PMCPMC6434516

11. Filmer D, Pritchett L. Child mortality and public spending on health: how much does money matter? http://documents.worldbank.org/curated/en/885941468741341071/Child-mortality-and-public-spending-onhealth-how-much-does-money-matter. Accessed 17 Jan 2022.

12. Roy S, Khatun T. Effect of adolescent female fertility and healthcare spending on maternal and neonatal mortality in low resource setting of South Asia. Health economics review. 2022;12(1):47. Epub 2022/09/18. doi: 10.1186/s13561-022-00395-7. PubMed PMID: 36115901; PubMed Central PMCID: PMCPMC9482740.

13. Moler-Zapata S, Kreif N, Ochalek J, Mirelman AJ, Nadjib M, Suhrcke M. Estimating the Health Effects of Expansions in Health Expenditure in Indonesia: A Dynamic Panel Data Approach. Applied health economics and health policy. 2022;20(6):881–91. Epub 2022/08/24. doi: 10.1007/s40258-022-00752-x. PubMed PMID: 35997895.

14. Abbasi BN, Sohail A. Ramification of healthcare expenditure on morbidity rates and life expectancy in the association of southeast asian nations countries: A dynamic panel threshold analysis. The International journal of health planning and management. 2022;37(6):3218–37. Epub 2022/08/20. doi: 10.1002/hpm.3551. PubMed PMID: 35983663.

15. Owusu PA, Sarkodie SA, Pedersen PA. Relationship between mortality and health care expenditure: Sustainable assessment of health care system. PloS one. 2021;16(2):e0247413. Epub 2021/02/25. doi: 10.1371/journal.pone.0247413. PubMed PMID: 33626059; PubMed Central PMCID: PMCPMC7904168.

16. Makela SM, Dandona R, Dilip TR, Dandona L. Social sector expenditure and child mortality in India: a state-level analysis from 1997 to 2009. PloS one. 2013;8(2):e56285. Epub 2013/02/15. doi: 10.1371/journal.pone.0056285. PubMed PMID: 23409166; PubMed Central PMCID: PMCPMC3567038.

17. Logarajan RD, Nor NM, Sirag A, Said R, Ibrahim S. The Impact of Public, Private, and Out-of-Pocket Health Expenditures on Under-Five Mortality in Malaysia. Healthcare (Basel, Switzerland). 2022;10(3). Epub 2022/03/26. doi: 10.3390/healthcare10030589. PubMed PMID: 35327065; PubMed Central PMCID: PMCPMC8953126.

18. Maruthappu M, Ng KY, Williams C, Atun R, Zeltner T. Government health care spending and child mortality. Pediatrics. 2015;135(4):e887–94. Epub 2015/03/04. doi: 10.1542/peds.2014-1600. PubMed PMID: 25733755.

19. Rana RH, Alam K, Gow J. Health expenditure, child and maternal mortality nexus: a comparative global analysis. BMC international health and human rights. 2018;18(1):29. Epub 2018/07/18. doi: 10.1186/s12914-018-0167-1. PubMed PMID: 30012137; PubMed Central PMCID: PMCPMC6048901.

20. Dhrifi A. Public Health Expenditure and Child Mortality: Does Institutional Quality Matter? J Knowl Econ. 2020;11:692–706.

21. WHO. Data explorer. Global health expenditure database. http://apps.who.int/nha/database/Select/Indicators/en. Accessed 11 September 2023.

22. Global, regional, and national progress towards Sustainable Development Goal 3.2 for neonatal and child health: all-cause and cause-specific mortality findings from the Global Burden of Disease Study 2019. Lancet (London, England). 2021;398(10303):870–905. Epub 2021/08/21. doi: 10.1016/s0140-6736(21)01207-1. PubMed PMID: 34416195; PubMed Central PMCID: PMCPMC8429803

23. Mohanty RK, Behera DK. Heterogeneity in health funding and disparities in health outcome: a comparison between high focus and non-high focus states in India. Cost effectiveness and resource allocation : C/E. 2023;21(1):44. Epub 2023/07/18. doi: 10.1186/s12962-023-00451-x. PubMed PMID: 37461113; PubMed Central PMCID: PMCPMC10351161.

24. Kiross GT, Chojenta C, Barker D, Loxton D. The effects of health expenditure on infant mortality in sub-Saharan Africa: evidence from panel data analysis. Health economics review. 2020;10(1):5. Epub 2020/03/08. doi: 10.1186/s13561-020-00262-3. PubMed PMID: 32144576; PubMed Central PMCID: PMCPMC7060592.

25. Qin VM, Hone T, Millett C, Moreno-Serra R, McPake B, Atun R, et al. The impact of user charges on health outcomes in low-income and middle-income countries: a systematic review. BMJ global health. 2018;3(Suppl 3):e001087. Epub 2019/02/23. doi: 10.1136/bmjgh-2018-001087. PubMed PMID: 30792908; PubMed Central PMCID: PMCPMC6350744.

26. Bein MA, Unlucan D, Olowu G, Kalifa W. Healthcare spending and health outcomes: evidence from selected East African countries. African health sciences. 2017;17(1):247–54. Epub 2017/10/14. doi: 10.4314/ahs.v17i1.30. PubMed PMID: 29026399; PubMed Central PMCID: PMCPMC5636241.

27. Garcia LP, Schneider IJC, de Oliveira C, Traebert E, Traebert J. What is the impact of national public expenditure and its allocation on neonatal and child mortality? A machine learning analysis. BMC public health. 2023;23(1):793. Epub 2023/04/29. doi: 10.1186/s12889-023-15683-y. PubMed PMID: 37118765; PubMed Central PMCID: PMCPMC10141942.

28. Li J, Yuan B. Understanding the effectiveness of government health expenditure in improving health equity: Preliminary evidence from global health expenditure and child mortality rate. The International journal of health planning and management. 2019;34(4):e1968–e79. Epub 2019/06/22. doi: 10.1002/hpm.2837. PubMed PMID: 31222802.

29. Rasella D, Basu S, Hone T, Paes-Sousa R, Ocké-Reis CO, Millett C. Child morbidity and mortality associated with alternative policy responses to the economic crisis in Brazil: A nationwide microsimulation study. PLoS medicine. 2018;15(5):e1002570. Epub 2018/05/23. doi: 10.1371/journal.pmed.1002570. PubMed PMID: 29787574; PubMed Central PMCID: PMCPMC5963760.

